# Eliciting and prioritising determinants of improved care in Multiple Long Term Health Conditions (MLTC): A modified online Delphi study

**DOI:** 10.1101/2023.03.19.23287406

**Authors:** Glenn Simpson, Beth Stuart, Marisza Hijryana, Ralph Kwame Akyea, Jonathan Stokes, Jon Gibson, Karen Jones, Leanne Morrison, Miriam Santer, Michael Boniface, Zlatko Zlatev, Andrew Farmer, Hajira Dambha-Miller

**Author notes:** **Corresponding author** Dr Glenn Simpson, Primary Care Research Centre, The University of Southampton, Aldermoor Close, SO16 5ST.

## Abstract

**Introduction:** Multiple Long Term Conditions (MLTC) are a major challenge to health and social care systems around the world. There is limited research exploring the wider contextual determinants that are important to improving care for people living with MLTC. In this study, we aimed to elicit and prioritise determinants of improved care in people with MLTC.

**Methods:** A three round online Delphi study was conducted in England with health and social care professionals, data scientists, researchers, people living with MLTC and their carers.

**Results:** Our findings suggest a care system which is still predominantly single condition focused. ‘Person-centred and holistic care’ and ‘coordinated and joined up care’, were highly rated determinants in relation to improved care for MLTC. We further identified a number of non-medical determinants that are important to providing holistic care for MLTC.

**Conclusions:** Further progress towards a holistic and patient centred model is needed to ensure that care more effectively addresses the complex range of medical and non-medical needs of people living with MLTC. This requires a move from a single condition focused biomedical model to a person-based biopsychosocial approach, which has yet to be achieved.

## Introduction

Multiple Long Term Conditions (MLTC) occur when an individual has two or more co-existing and enduring health conditions [1]. The prevalence of MLTC is an immediate priority and a long term challenge to health and social care systems around the world owing to the projected rise in the number of people aged over 65 years with these conditions [2,3]. This is not only the case in high income countries where MLTC is “considered the norm, not the exception” [4, p.5], but prevalence is also increasing rapidly in lower income states. Globally, this will impose significant challenges at individual, care system and wider societal levels, as MLTC requires more intensive and complex care and is associated with higher levels of service usage and expenditure, increased sickness absence rates and a greater risk of social vulnerability (including homelessness, unemployment and poverty) [4-6].

MLTC is also a challenge for professionals providing care for this cohort, as health and social care systems (including clinical training and clinical practice guidelines) [7], are primarily based on a single disease paradigm [8,9]. This is problematic, as the existing focus on single disease approaches based on a highly specialised care and treatment regime [8] may not comprehensively address the complex range of care needs which are a feature of MLTC [10]. A recent scoping review [10] found evidence that experiences of living with and seeking care for MLTC are not only shaped by medical factors and the quality of clinical care, but also by wider non-medical determinants (also referred to as social determinants) of health [11] such as housing conditions, income and employment, and the quality of the local environments (access to green space, air quality, and neighbourhood crime levels and social cohesion).

These non-medical determinants interact with each other and also with medical factors to determine an individual’s probability of developing MLTC and their subsequent deterioration in health [6, 9-11]. Whilst there are potentially a limitless number of medical and non-medical determinants that could be examined across all dimensions of care, it is unclear which of these determinants should be prioritised to deliver effective interventions for MLTC [10].

Currently, there is a limited body of research exploring the full range of determinants which are critical to improving care in MLTC [9, 12]. Given these limitations in the current evidence-base, we elicited opinions from health and social care professionals, data scientists, researchers, people living with MLTC and their carers, in order to prioritise wider determinants of improved care in MLTC, with a view to informing efforts to develop new models of care that might address the holistic needs of people with MLTC [2, 13].

## Method

### Theoretical framework

The Sustainable intEgrated care models for multi-morbidity: delivery, FInancing and performancE (SELFIE) framework was used to conceptually inform our research design, methods and interpretations [14]. SELFIE was developed as a framework to support the design, development, assessment and delivery of programmes of integrated care for individuals with MLTC [14]. Theoretically, the framework is constructed around the holistic care needs of the individual with MLTC as experienced within their immediate environment. In this context, the individual with MLTC is considered centrally, with surrounding and radiating concepts grouped into six adapted World Health Organisation components “used to describe, understand, and compare different health systems” (i.e.: 1. Service delivery; 2. Leadership and governance; 3. Workforce; 4. Financing; 5. Technologies and medical products; 6. Information and research) [14, p.5]. Within each component, concepts are further divided into three spatial scales (i.e. micro, meso and macro) that identify and prioritise the levels at which interventions, policy action and resources can be most effectively delivered by care providers to manage the care needs of those with MLTC.

### Study Design

The Delphi technique is a well-established formal “group facilitation” [15, p.1008] method to gather anonymous opinions from a panel of experts with the aim of achieving consensus [16-17] and is often used in research to determine priorities [18]. This method was chosen for its flexibility, allowing us to elicit and prioritise a broad range of views on wider determinants relevant to improved care in MLTC.

### Recruitment and sampling

There is not a standard figure in the literature for the number of participants required to conduct a Delphi study, although Vogel et al. [19, p.2576] suggest that a “minimum of 12 respondents is generally sufficient to enable consensus.” Using this as a baseline number and taking into account the potential for participant ‘drop-out’ over the duration of the three rounds of the study [20], we aimed to recruit at least 20 participants. Purposive sampling was used to ensure a diverse range of participants in terms of backgrounds and experiences. We recruited via online forums, social media, the websites of relevant service provider organisations such as local government adult social care and voluntary sector groups. We also used snowball sampling whereby respondents who had agreed to be involved in the study were asked to recommend other participants to join the Delphi panel [21]. All participants provided written consent to participate in the study.

### Survey development

The study was informed by the research question, literature review, public and patient involvement and input from the wider study team [17]. The study was piloted before being deployed. Each round included data collection of socio-demographic characteristics.

Round 1 asked a single open question: What medical and non-medical factors are key to improving the care of people with multiple long term conditions (e.g. medical, social, environmental, economic or other factors)? In response, participants were asked to provide up to 20 items.

The purpose of round 1 was to encourage item generation by the participants with the subsequent rounds 2 and 3 focused on item prioritisation using a seven-point Likert Scale (ranging from: 1= not at all important, to 7= extremely important) [22]. Each round included a free-text box to allow respondents to add additional comments to elaborate further on their responses. Each stage of the Delphi process is set out in Figure 1.

**Figure 1:**
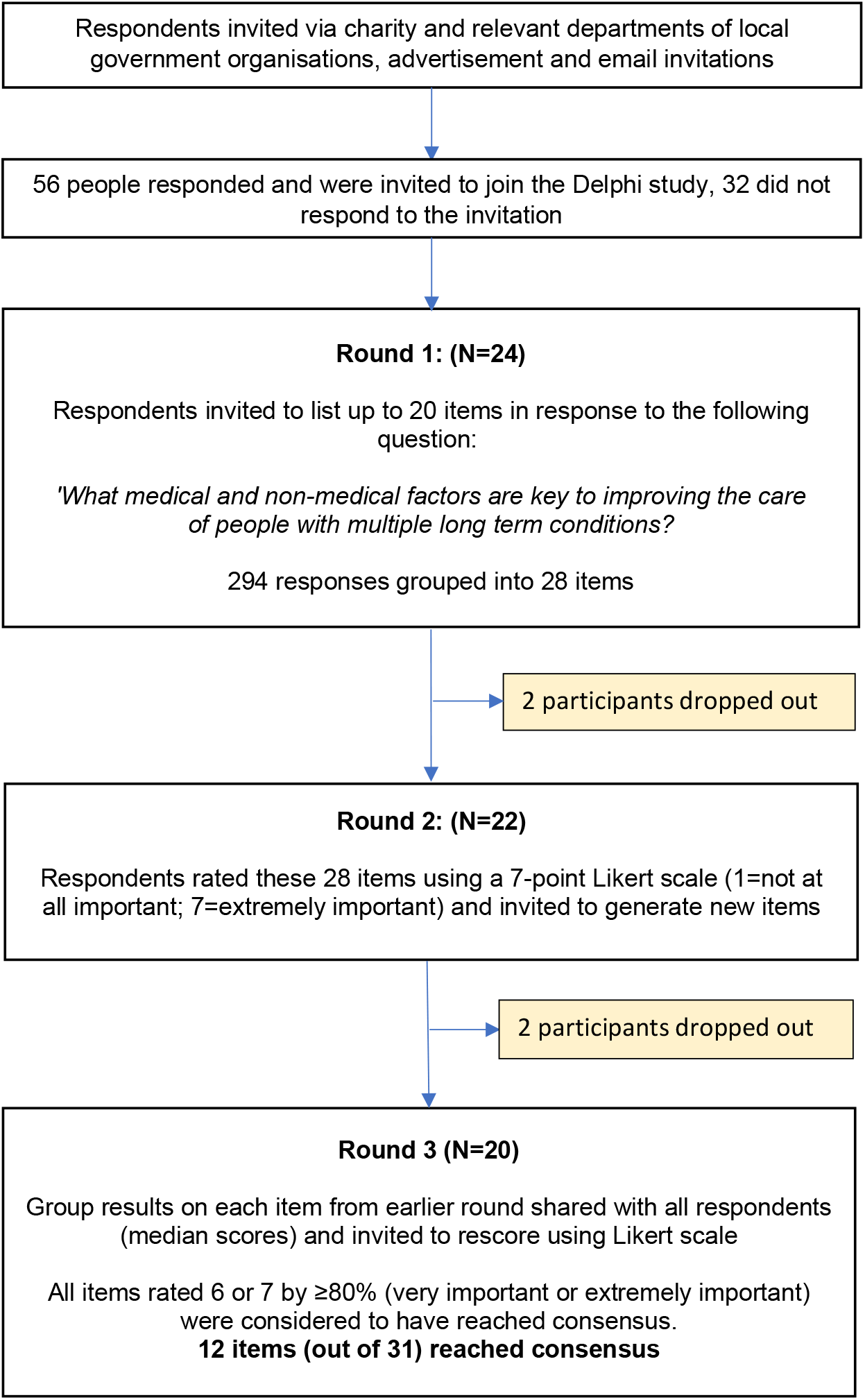
Flow chart summarising each stage of the Delphi process.

### Data collection

Data collection was conducted between February and April 2022. A web-link to access the Delphi study was emailed to participants. An online format was used due to national restrictions imposed by the COVID-19 pandemic that limited in-person meetings and to facilitate greater geographical diversity of participants. Those who expressed interest in the study were sent an online study information sheet. Non-responses were followed up with up to three email reminders sent to respondents. Following these reminders, participants were classified as ‘drop-outs’ if no response was received within a month [23]. No participation incentives were used. The Delphi study lasted three rounds as sufficient consensus was achieved at this stage.

The data collected was transferred to the NVivo 12 qualitative analysis platform for collation and analysis. Consensus was defined by a priori criteria of 80% or more responses falling within the top two categories on the Likert Scale (6: very important or 7: extremely important) [24].

## Results

### Participants

All 56 respondents who expressed interest were invited to participate in the study. Of these 24 (43%) commenced round one and 20 of these initial respondents completed round three (83% follow-up). The flow of participants through each round is summarised in Figure 1.

An equal number of males and females completed the study, and the majority of respondents (83%) were from White British backgrounds. Most respondents resided in England (88%). All participants were over 18 years old. Those aged 40-49 were the single largest category of respondents (33%), although only one respondent was under 30 years old and three (13%) were in the oldest age range of 60-69 years, a period during the life course which is most commonly associated with MLTC prevalence. Respondents from care service providers and healthcare backgrounds were the two largest groups on the panel. A summary of the socio-demographic and occupational characteristics of respondents (who initially participated in round 1) are shown in table 1 below.

**Table 1:** Socio-demographic and occupational characteristics of respondents.

| **Respondent Characteristics** | **Results, n (%)** |
| --- | --- |
| **Gender** |  |
| Male | 12 (50) |
| Female | 12 (50) |
| **Age** |  |
| 18-29 | 1 (4) |
| 30-39 | 6 (25) |
| 40-49 | 8 (33) |
| 50-59 | 6 (25) |
| 60-69 | 3 (13) |
| **Ethnicity** |  |
| British | 20 (83) |
| Any other White background | 2 (8) |
| Chinese | 1 (4) |
| Mixed (White & Black Caribbean) | 1 (4) |
| **UK nation of residence** |  |
| England | 21 (88) |
| Wales | 1 (4) |
| Scotland | 2 (8) |
| **Current role/position** |  |
| Healthcare professionals | 5 (21) |
| Social care professionals | 3 (13) |
| Health data scientists | 2 (8) |
| Researchers | 3 (13) |
| Care service providers | 6 (25) |
| Voluntary sector | 3 (13) |
| Adults with MLTC | 2 (8) |

### Round 1

A total of 294 individual item responses were received in round 1. These items were grouped into related categories and labelled. The analysis was guided and structured using the SELFIE theoretical framework. Specifically, this involved the lead researcher systematically mapping items onto the SELFIE conceptual categories listed on Leijten et al’s [14] diagrammatic model of integrated care for MLTC, a process that was repeated for additional items suggested in subsequent rounds (Table 2).

**Table 2:** Delphi determinants mapped onto SELFIE framework. Those Delphi determinants highlighted in red aligned with SELFIE framework

| Core of the framework | LEVELS OF INTEGRATED CARE FOR MULTIMORBIDITY |  |  | Six key components of a well-functioning health system |
| --- | --- | --- | --- | --- |
|  | Micro-scale | Meso-scale | Macro-scale |  |
| **Holistic understanding of the individual with multimorbidity:**<br>Health<br>Wellbeing<br>Capabilities<br>Self-management<br>Needs<br>Preferences | Person-centred | Organisational and structural integration | Market regulation | Service Delivery |
|  | Proactive | Continuous quality improvement system | Policies to integrate care across organisations and sectors |  |
|  | Tailored |  | Service availability and access |  |
|  | Self-management |  |  |  |
|  | Informal caregiver involvement |  |  |  |
|  | Treatment interaction |  |  |  |
|  | Continuity |  |  |  |
|  | Coordination tailored to complexity | Performance-based management | Policy and action plans on chronic diseases and multimorbidity | Leadership and Governance |
|  | Shared decision-making | Supportive leadership | Political commitment |  |
|  | Individualised care planning | Clear accountability |  |  |
|  |  | Performance-based management |  |  |
|  |  | Culture of shared vision, ambition, values |  |  |
| **Environment:**<br>Housing<br>Social network<br>Transport<br>Community<br>Welfare services<br>Financing | Multidisciplinary team | Continuous [professional] development | Educational and workforce planning | **Workforce** |
|  | Named coordinator | Informal caregiver support | Workforce and demography match |  |
|  | Core Group | New professional roles |  |  |
|  | Coverage and reimbursement | Incentives to collaborate | Equity and access | **Financing** |
|  | Out of pocket costs | Risk adjustments | Financial system for health and social care |  |
|  | Financial incentives | Shared savings | Stimulating investments in innovative care models |  |
|  |  | Secured budgets |  |  |
|  |  | Business case |  |  |
|  | EMR and patient portals | Shared information systems | Policies fostering technological innovation | **Technologies and medical products** |
|  | E-health tools | Interoperable systems | Access to technologies and medical products |  |
|  | Assistive technologies |  |  |  |
|  | Remote monitoring |  |  |  |
|  | Individual data | Data ownership and protection | Access to information | **Information and research** |
|  | Individual risk prediction | Innovative research [methods] | Policies that stimulate research in integrated care and multimorbidity |  |
|  |  | Risk stratification | Privacy and data protection legislation |  |

The mapping process identified items that did not clearly align with the SELFIE categories. The relevance of ‘non-aligned’ items and whether they should be included in subsequent rounds of the study were reviewed against the existing MLTC literature by the lead researcher, who also discussed the relevance of these items with the two other researchers involved in the Delphi study. This provided an additional layer of independent scrutiny over the conclusions reached about the relevance of individual items. Items not aligned to the SELFIE framework but considered relevant to the study’s aims were included in subsequent rounds for deliberation (these are marked on Table 3).

**Table 3:** Participant Responses Round 1.

| Item Number | Suggested priority determinants of improved care in MLTC |
| --- | --- |
| 1 | Person-centred and holistic care |
| 2 | Continuity of care |
| 3 | Early intervention and preventative approach |
| 4 | Inclusive/Equitable service provision |
| 5 | Access to social care, community-based services and other provision |
| 6 | Service availability in terms of timeliness and geography |
| 7 | Training in multiple long term conditions for healthcare and social care professionals |
| 8 | Coordinated and joined up care |
| 9 | Data sharing across care services and linked electronic health records |
| 10 | Thorough assessment of health and social care needs |
| 11 | Supporting self-management of conditions |
| 12 | Symptom management and monitoring |
| 13 | Access to information for patients and carers and where to go for support |
| 14 | Use of technologies to support individuals at home |
| 15 | Prescribing and medication management |
| 16 | Enhanced support from family and other informal carers |
| 17 | Support provided to informal carers |
| 18 | Support network |
| 19 | Addressing loneliness and social isolation |
| 20 | Recognition of and support with lifestyle factors |
| 21 | Environmental factors and wider social determinant of health |
| 22 | Finance/financial assistance |
| 23 | Housing/accommodation that meets individual's needs |
| 24 | Increasing knowledge and awareness of multiple long term conditions among, patients, carers and people in the wider community ~ |
| 25 | Support for mental health and emotional wellbeing ~ |
| 26 | Able to engage in meaningful activities and social participation ~ |
| 27 | Support with daily living and independent living ~ |
| 28 | Work issues and training for people living with multiple long term conditions ~ |
| ~ Items not aligned with the SELFIE Framework |  |

The mapping process synthesised the individual response items into 28 categories, defined as priority determinants (see Table 3). These priority determinants of improved MLTC were then presented to respondents in round 2.

### Round 2

Round 2 allowed respondents to rate the 28 determinants generated in round 1 (Table 4). Median scores were then calculated for each determinant based on the average of the Likert Scale rating. A clear hierarchy of prioritisation emerged, with the highest median scores given for systems-wide determinants ‘person-centred and holistic care’ (ranked 1) and ‘co-ordinated and joined up care’ (ranked 2). Determinants focused on care delivery, ‘thorough assessment of health and social care needs’ and ‘support for mental health and emotional wellbeing’, were ranked three and four respectively. The social determinant of health, ‘housing/accommodation that meets individual’s needs’, also received a high rating, ranked five.

**Table 4:** Median Ratings of Priority Determinants in Round 2.

| Number | Priority Determinants | Median |
| --- | --- | --- |
| 1 | Person-centred and holistic care | 7 |
| 2 | Coordinated and joined up care | 7 |
| 3 | Thorough assessment of health and social care needs | 7 |
| 4 | Support for mental health and emotional wellbeing | 7 |
| 5 | Housing/accommodation that meets individual's needs | 6.5 |
| 6 | Continuity of care | 6 |
| 7 | Early intervention and preventative approach | 6 |
| 8 | Inclusive/Equitable service provision | 6 |
| 9 | Access to social care, community-based services and other provision | 6 |
| 10 | Service availability in terms of timeliness and geography | 6 |
| 11 | Training in multiple long term conditions for healthcare and social care professionals | 6 |
| 12 | Data sharing across care services and linked electronic health records | 6 |
| 13 | Supporting self-management of conditions | 6 |
| 14 | Symptom management and monitoring | 6 |
| 15 | Access to information for patients and carers and where to go for support | 6 |
| 16 | Prescribing and medication management | 6 |
| 17 | Addressing loneliness and social isolation | 6 |
| 18 | Support provided to informal carers | 6 |
| 19 | Environmental factors and wider social determinant of health | 6 |
| 20 | Finance/financial assistance | 6 |
| 21 | Able to engage in meaningful activities and social participation | 6 |
| 22 | Support with daily living and independent living | 6 |
| 23 | Increasing knowledge and awareness of multiple long term conditions among patients, carers and people in the wider community | 5.5 |
| 24 | Enhanced support from family and other informal carers | 5.5 |
| 25 | Recognition of and support with lifestyle factors | 5.5 |
| 26 | Work issues and training for people living with multiple long term conditions | 5 |
| 27 | Use of technologies to support individuals at home | 5 |
| 28 | Support network | 5 |

Respondents were also able to suggest new determinants to add to the round 1 list. Three respondents proposed additional determinants for consideration in round 3: 1) ‘support for children of people living with multiple long term conditions’; 2) ‘more intervention from private care providers’; 3) ‘flexible systems in place to support people (before ‘cliff-edge’ moments when their conditions or circumstances change)’.

Three respondents added comments in the free text box, which were concerned with two themes: firstly, it was challenging to provide a rating, as the relative importance of each determinant depended on the personal circumstances of each individual and their specific care needs; and second, most determinants were interconnected, making it difficult to discriminate between their relative importance.

### Round 3

Respondents ‘re-prioritised’ the 28 determinants from earlier rounds after considering the group responses and also the three additional suggestions generated from the free-text responses in round 2. Respondents rated 12 out of 31 determinants as very important (6 or 7 on the Likert scale), thus reaching consensus as set out in our methods (Table 5).

**Table 5:** Final Consensus Results from Round 3.

| No. | Priority Determinants | Median score | Numbers (%) of participants who rated 6 (very important) or 7 (extremely important) N=20 | Consensus level and priority rating of factors for addressing care need in MLTC | Ranking position |
| --- | --- | --- | --- | --- | --- |
| 1. | Person-centred and holistic care | 7 | 20 (100) | Very high level of consensus/Very high priority | 1 |
| 2. | Coordinated and joined up care | 7 | 19 (95) | Very high level of consensus/Very high priority | 2 |
| 3. | Flexible systems in place to support people (before 'cliff-edge' moments when their conditions or circumstances change)*^ | 6 | 18 (90) | Very high level of consensus/Very high priority | 3 |
| 4. | Support for mental health and emotional wellbeing* | 6 | 17 (85) | High level of consensus/High priority | 4 |
| 5. | Data sharing across care services and linked electronic health records | 6 | 17 (85) | High level of consensus/High priority | 4 |
| 6. | Training in multiple long term conditions for healthcare and social care professionals | 6 | 17 (85) | High level of consensus/High priority | 4 |
| 7. | Access to social care, community-based services and other provision | 6 | 17 (85) | High level of consensus/High priority | 4 |
| 8. | Thorough assessment of health and social care needs | 6 | 17 (85) | High level of consensus/High priority | 4 |
| 9. | **Early intervention and preventative approach** | 6 | **17 (85)** | High level of consensus/High priority | **4** |
| 10. | **Continuity of care** | 6 | **17 (85)** | High level of consensus/High priority | **4** |
| 11. | **Service availability in terms of timeliness and geography** | 6 | **17 (85)** | High level of consensus/High priority | **4** |
| 12. | **Supporting self-management of conditions** | 6 | **16 (80)** | High level of consensus/High priority | **5** |
| 13. | Symptom management and monitoring | 6 | 15 (75) | Fairly high level of consensus below threshold | **6** |
| 14. | Prescribing and medication management | 6 | 14 (70) | Fairly level of consensus below threshold | **7** |
| 15. | Inclusive/Equitable service provision | 6 | 14 (70) | Fairly high level of consensus below threshold | **7** |
| 16. | Support with daily living and independent living* | 6 | 14 (70) | Fairly high level of consensus below threshold | **7** |
| 17. | Addressing loneliness and social isolation | 6 | 14 (70) | Fairly high level of consensus below threshold | **7** |
| 18. | Finance/financial assistance | 6 | 13 (65) | Some level of consensus below threshold | **8** |
| 19. | Support provided to informal carers | 6 | 13 (65) | Some level of consensus below threshold | **8** |
| 20. | Access to information for patients and carers and where to go for support | 6 | 12 (60) | Some level of consensus below threshold | **9** |
| 21. | Environmental factors and wider social determinants of health | 6 | 12 (60) | Some level of consensus below threshold | **9** |
| 22. | Able to engage in meaningful activities and social participation* | 6 | 12 (60) | Some level of consensus below threshold | **9** |
| 23. | Recognition of and support with lifestyle factors | 6 | 11 (55) | Some level of consensus below threshold | **10** |
| 24. | Support for children of people living with multiple long term conditions*^ | 6 | 11 (55) | Some level of consensus below threshold | **10** |
| 25. | Housing/accommodation that meets individual's needs | 6 | 11 (55) | Some level of consensus below threshold | **10** |
| 26. | Work issues and training for people living with multiple long term conditions* | 5 | 9 (45) | No consensus | **11** |
| 27. | Enhanced support from family and other informal carers | 5 | 8 (40) | No consensus | **12** |
| 28. | Increasing knowledge and awareness of multiple long term conditions among patients, carers and people in the wider community* | 5 | 8 (40) | No consensus | **12** |
| 29. | Support network | 5 | 7 (35) | No consensus | **13** |
| 30. | More intervention from private care providers *^ | 5 | 6 (30) | No consensus | **14** |
| 31. | Use of technologies to support individuals at home | 5 | 4 (20) | No consensus | **15** |
| Factors reached consensus threshold if ≥80% rated, either 6 or 7 on Likert Scale, are shown in **bold**.<br>*Factors not aligned with the SELFIE Framework.<br>^Additional determinants suggested by respondents in round 2. |  |  |  |  |  |

Consensus was not reached in the other 19 determinants, although 12 of these achieved some degree of consensus, albeit below our pre-determined threshold. For the remaining six determinants, there was no consensus among respondents.

There were three areas of very high consensus. All participants (100%) agreed that ‘person-centred and holistic care’ was extremely important. Nineteen (95%) and 18 (90%) out of 20 participants rated ‘coordinated and joined-up care’ and ‘flexible systems in place to support people (before ‘cliff-edge’ moments when their conditions or circumstances change)’, as extremely important (Table 5).

Eight other determinants equally achieved a high level of consensus, with 17 out 20 (85%) of participants rating these as very important (1. Support for mental health and emotional wellbeing; 2. Data sharing across care services and linked electronic health records; 3. Training in multiple long term conditions for healthcare and social care professionals; 4. Access to social care, community-based services and other provision; 5. Thorough assessment of health and social care needs; 6. Early intervention and preventative approach; 7. Continuity of care; 8. Service availability in terms of timeliness and geography). One other determinant, supporting self-management of conditions reached the consensus threshold, with 16 out of 20 respondents (80%) rating this determinant as very important.

Comparing these results with round 2 rankings, it is evident that over each round of the study, ‘person-centred and holistic care’ and ‘coordinated and joined up care’ consistently achieved very high levels of consensus, indicating these were considered by respondents to be the highest priority determinants for improving care in MLTC. Further, in the final round of deliberation, the additional determinant suggested in round 2, ‘flexible systems in place to support people (before ‘cliff-edge’ moments when their conditions or circumstances change), emerged as the third highest rated determinant. Other determinants suggested in round 2 (‘more intervention from private care providers’ and ‘support for children of people living with multiple long term conditions’) did not achieve the consensus threshold. A notable change in ranking was observed in relation to the determinant ‘housing/accommodation that meets individual’s needs’, which respondents rated in the top five of priority determinants in round 2 but failed to reach the consensus threshold in round 3. Apart from the examples discussed above, respondents in round 3 did not significantly change their previous rating of each determinant from round 2, indicating a level of consistency in the prioritisation responses throughout the study.

Amongst eight respondents who left various comments in the free-text box, two mentioned how they changed their responses in round 3. One respondent changed some responses based on their reflections of the wider Delphi group results while another mentioned that their re-rating was not affected by the group results. Five of the eight respondents left comments, including: 1) all determinants were important and that they overlapped to some extent; 2) most of the determinants were significant for different people at different stages of their health trajectory.

### MLTC priority determinants and the SELFIE Framework

The significant majority of MLTC determinants (i.e. 24 out of 31 or 77%) (Table 5) identified by respondents aligned conceptually with one or more of the SELFIE framework categories [14]. Comparing these priority determinants to the six health systems components of the framework [14], the majority were weighted towards the ‘Service delivery’ component of the framework, and to a lesser extent the ‘Technologies and medical products’, ‘Workforce’ and ‘Financing’ components.

Service delivery responses were focused on the need for tailored, personalised services including treatment interaction (between the patient and care providers), patient self-management, the need for multi-disciplinary teams and ‘named’ care co-ordinators, informal carer involvement in decisions made about care options and maintaining continuity of care. Additionally, there was an emphasis on policies ensuring that services were both available and accessible in local communities, especially for patients with restricted mobility or limited transport options.

Technologies and medical product responses prioritised the need for shared information and monitoring systems across organisations. These included technologies which enhance information sharing and access relevant to frontline practitioners and patients (especially in relation to self-management of conditions), such as electronic medical records, patient portals, e-health tools and assistive technologies.

Workforce responses focused on structures and co-ordination roles that facilitate integrated care across organisational boundaries, in particular multidisciplinary teams and a named care co-ordinator or case manager. Further, there was an emphasis on continuous professional development in MLTC for care professionals, and provision of informal caregiver support, a cohort who often play a critical role in the care process [14].

In relation to the Financing component of the framework, aspects of financial support for those with MLTC were emphasised. These included: “coverage and reimbursement of the interventions included in person-centred integrated care programmes [which] need to be generous enough to ensure equity in financial access for those who need them”; and out of pocket costs, which “may influence access, [non]adherence, and how and which care is used” [14, p.17]. In addition, financial incentives were prioritised which can be “used to motivate persons with multi-morbidity to participate in and adhere to integrated care programmes” [14, p.17], as well as the need for policy-makers to ensure equity and equal access to services for people with MLTC, especially those from lower socio-economic backgrounds.

Relating the priority determinants to the SELFIE scales of integration, the majority were located at the micro scale (n=9) of the framework. This is the scale concerned with the frontline delivery of care and other support to the individual. This indicates a prioritisation among respondents focused on promoting integrated care for those living with MLTC, which is personalised to individual care needs across all components of the health system.

Five determinants aligned to the meso scale and two to the macro scale. The meso and macro scales in the SELFIE framework are concerned with the policy, planning, organisational and operational components that underpin effective delivery of integrated frontline care provision. The determinants prioritised by respondents mainly emphasised the need for co-ordinated services, processes and structures which facilitate joined up care, in particular shared information systems, organisational integration, interoperable systems and specific training and professional development for care practitioners in MLTC.

Virtually all determinants were aligned to the core of the SELFIE model (which in relation to the hierarchy of scales in the SELFIE model can be understood as the individual scale), that is the holistic understanding of an individual’s specific care needs and capabilities to sustain and manage their care. All of the environment elements at the core of the framework (which also encompass wider social determinants of health) aligned to the priority determinants identified by our study.

A number of priority determinants (7 of 31 or 22.5%) identified were not closely aligned with or did not feature prominently in the SELFIE framework, although some of these were considered by respondents to be relevant to established MLTC (Table 5). The determinant ‘support for mental health and emotional wellbeing’ consistently received a high ranking by participants throughout the study. Two other determinants, which did not reach the consensus threshold, were nevertheless considered to be ‘very important’ in relation to improving care in MLTC by 70% of respondents (‘Support with daily living and independent living’) and 60% (‘Able to engage in meaningful activities and social participation’) respectively. Interestingly, these determinants highlight the importance of a holistic approach to care in MLTC encompassing an individual’s daily social care needs, addressing their psychological and mental wellbeing and wider social needs.

## Discussion

In this study, we used the Delphi method to elicit and prioritise determinants of improved care in MLTC. A clear hierarchy of the key priority determinants were identified by our diverse panel of patients, carers, health and social care professionals, data scientists, and researchers in MLTC. There was a near unanimity of consensus among respondents that the primary priority determinants of improved care in MLTC were ‘person-centred and holistic care’ (ranked 1) and ‘coordinated and joined up care’ (ranked 2). Further, the determinant ‘flexible systems in place to support people (before ‘cliff-edge’ moments when their conditions or circumstances change)’ (ranked 3), also achieved a very high level of consensus among the panel. These priority determinants suggest that improving care for those living with MLTC requires a paradigm change in the current model of care and practice. In other words, there is a need for a more concerted move towards a holistic and individualised care model, built around the needs of the patient, which is organised and delivered by integrated care services to ensure continuity of care and seamless transitions in care between service providers. Similar findings have also been reported by a range of other studies examining improved care in MLTC [25, 2, 1, 9, 26, 27].

Other priority determinants for improved care which achieved consensus reflected the complex and diverse range of medical and non-medical care needs associated with MLTC and issues relating to access to services. These included support for mental health and wellbeing for those with MLTC, access to and availability of social services and other provision in local communities and comprehensive assessment of health and social care needs of individuals with MLTC. Supporting patients with self-management of conditions was also identified as a priority determinant, highlighting a need for a shift towards greater patient participation in their care, which is “an essential component of chronic disease management and secondary prevention” [28], as well as being an important dimension of the person-centred care agenda.

Whilst there was a strong focus on improved frontline care at the level of the individual, our findings also reflected an emphasis on macro factors relating to systems change and service reconfiguration. These included data sharing across care services, linked electronic health records, continuity of care and early intervention and preventative approaches [25], the latter also indicating a need to address upstream social and non-medical determinants which can significantly influence health trajectories and outcomes [29].

An interesting finding from our study was the identification of training in multiple long term conditions for healthcare and social care professionals as a priority determinant. This is an issue which has received relatively little attention in the literature [25] and may indicate that current training programmes for healthcare and social care professionals continue to be specialist and designed around treatment and care for single conditions, and as such fail to adequately provide care personnel with the requisite skills and knowledge that is specific to care for MLTC.

Overall, most of our principal findings are reflected in the wider literature [8] and perhaps are indicative of a care system in the UK that is still predominantly structured and delivered on basis of care for single conditions, with care practice and organisational cultures also similarly orientated. In such an institutional and operational form, the system likely struggles to address holistically, the multifaceted and complex care needs associated with MLTC [2]. Considered from a strategic policy perspective, the key priority determinants identified may be a recognition that efforts to advance person-centred care, which is holistic, integrated and addresses a range of interrelated medical and non-medical care needs of MLTC patients, continues to be slow, and implementation remains a major policy and practice challenge both in the UK and internationally [2, 26, 4, 8]. An important learning point for policy-makers from our study is that these are likely to continue to feature as critical determinants influencing care in MLTC until further significant progress is made towards redesigning care services and delivery around the needs of the individual rather than service providers [26,25]. In relation to care practice, our findings suggest that clinicians and care practitioners need to take further steps to ensure wider non-medical determinants and care needs are fully assessed and taken into consideration in care plans and subsequent delivery of care to those with MLTC [2, 10].

## Strengths and limitations

Based on our knowledge of the literature, this is one of the first studies to use the Delphi method to identify and prioritise those medical and non-medical determinants that are key to improving the care of people with MLTC. The main strength of our study is the diverse and balanced panel of respondents spread across the age range, including people living with MLTC and those providing informal and formal care. This provided a more comprehensive understanding of MLTC and determinants of importance. The online Delphi approach may have contributed to capturing a geographically diverse UK-wide sample, as well as contributing to the high response rate (83%), which is above most recommendations [30,31]. For this study, three rounds were found to be an appropriate duration, as the results obtained in the second and third rounds did not change significantly, indicating it was unlikely important additional data would have been generated by holding further rounds of deliberation. Extending the duration of the study may also have affected the response rate, potentially undermining the validity of the results [23].

However, as an online Delphi study, the absence of an interactive in-person discussion may have limited the richness of data collection as respondents were unable to engage in dialogue and exchange opinions with others on the panel regarding prioritisation of determinants. While our sample size was within the accepted range for Delphi studies [20], a relatively low number of respondents participated in the study (n=20), the significant majority of whom (83%) were White British, which may have limited the range of perspectives and diversity of experiences captured. Further, a greater number of study participants may have enabled the collection of a broader range of perspectives on the topic area and produced more data. We do not know why only 32 of the 56 respondents who initially expressed an interest in participating in the study, subsequently decided not to do so. However, it is important to note that our sample remained stable throughout, with little respondent ‘drop-out’ recorded, a factor which may have supported a consistency of approach in reaching a final consensus of opinion.

Ranking of the determinants is inherently a subjective process influenced by respondents’ lived experience, specific knowledge and expertise, as well as professional backgrounds and training, etc. In this respect, most of our study participants were care professionals or from other professional backgrounds, sample characteristics which may have biased the selection and weighting of priority determinants.

Additionally, as some respondents commented, it can be difficult to assign a weighting to individual determinants, as prioritisation can be dependent on a complex range of variables and risks such as a patient’s specific personal circumstances including socio-economic status, the environmental and geographical context in which they live, as well as the stage in their disease trajectory, etc. As some respondents indicated, prioritisation is also a challenge because determinants are often inextricably interrelated and interdependent, making it difficult to separate out and identify a hierarchy of importance in relation to improving care in MLTC.

## Conclusion

Our study highlights the complex range of interrelated medical and non-medical determinants influencing care in MLTC. Current understanding of and approaches to care in MLTC are still too focused on the medical dimensions of care and priorities of systems and service providers [25]. However, the care needs of those living with MLTC are often broader, encompassing biopsychosocial needs (e.g. mental health and emotional wellbeing) [2, 25] and social circumstances including the impact on health and wellbeing of the wider environment in which an individual is situated [10]. Our study supports the growing body of evidence that further progress towards a holistic and person-centred care model [2, 25, 32] is urgently required, which empowers people to be full and equal participants in their care decision-making to ensure that care delivery can be further optimised to address the diverse range of individual care needs often associated with MLTC [2, 25, 33]. This requires a cultural shift in contemporary healthcare and social care philosophy and practice [25], necessitating a move ‘from a single disease-focused biomedical model to a biopsychosocial’ approach [12], as well as further progress towards an integrated care system [2]. While some progress has been made in this direction in recent years [2], such a shift in approach may also need to be complemented by a greater emphasis and re-orientation towards preventative upstream approaches [34]. This will necessitate directing increased expenditure on preventative public health activities including support to those agencies (e.g. local government) responsible for social care and addressing wider social or non-medical determinants of health [29]. A recent study [6] highlighted the importance of targeting resources towards preventative activities in relation to MLTC, concluding that: ‘multimorbidity appears to be associated with per capita public health expenditure, and health-related quality of life with social care expenditure.’

Additionally, the implementation of new care governance and delivery models such as Integrated Care Systems (ICS) in England may also provide a mechanism for integrated delivery of tailored person-centred care [35]. ICS are partnership bodies consisting of care provider organisations and local authorities, the purpose of which is to plan and deliver joined up health and care services [36]. Whilst there are tentative signs that ICS may have potential to enhance care integration [36], the concept is a recent innovation and as a result it is too early to make an assessment as to whether this model will be able to deliver the integrated and personalised models of care which policy-makers envisage.

## Data Availability

All data produced in the present study are available upon reasonable request to the authors.

## Acknowledgements

We would like to thank all of the professional, patient and public contributors of our study. Without their participation, this work would not have been possible.

## Contributors

HDM and AF developed the research question and study design. HDM and GS drafted the paper. MH and GS contributed to data collection and analysis. All authors contributed to the survey design, analysis and critically revised the manuscript.

## Patient Consent

Informed written consent was obtained for all participants.

## Ethics approval

Ethical approval for the study was provided by the Faculty of Medicine Ethics Committee, University Hospital Southampton (reference number 67953).

## Provenance and peer review

Not commissioned; externally peer-reviewed.

